# Feasibility of converting Japanese oncology electronic medical records into the Observational Medical Outcomes Partnership Common Data Model and data quality assessment

**DOI:** 10.1101/2025.06.13.25329609

**Authors:** Yoshihiro Aoyagi, Suzue Terao, Baba Masahiro, Keiichi Nomura, Yuuya Ikeda, Akihiro Sato

## Abstract

The potential of utilizing Japanese electronic medical record (EMR) data in global observational research is significant because of high EMR adoption and universal health insurance. However, a few studies have addressed the conversion of Japanese EMR data to the Observational Medical Outcomes Partnership Common Data Model (OMOP CDM) standard, which regulates EMR data for global observational research. In this study, we investigated the feasibility of converting Japanese oncology EMR data to the OMOP CDM and applying the Observational Health Data Sciences and Informatics (OHDSI) tools for analysis. We focused on data from the National Cancer Center Hospital East, encompassing 8,447 patients with breast cancer between January 2015 and November 2023. The main objectives included vocabulary standardization and data structure standardization. The anonymized dataset included clinical information such as patient demographics, diagnoses, treatments, and laboratory results. A total of 3,697 unique disease names, 987 specimen test result terms, and 1,144 drug terms were successfully mapped to OMOP CDM standards, with IC-10 terms showing the highest success rate for disease names. A total of 90% of clinical terms were successfully mapped to OMOP CDM standards, with 80% of source data fully integrated. However, only 32 surgical terms were identified. The feasibility of converting EMR data to OMOP CDM was evaluated by mapping source terms, comparing local raw datasets, and conducting a comprehensive quality assessment using a Data Quality Dashboard. A total of 1,991 validation checks were performed to evaluate the validity of data, suitability, and completeness. The results revealed 24 checks flagged as FAIL or ERROR, with the most frequent issues in the measurement table (10 errors). Despite these issues, the conversion process demonstrated high feasibility. Overall, this study positions Japan as a key player in international observational oncology research, enhancing the global understanding of treatment effectiveness and patient outcomes in real-world settings.

## Introduction

Real-world data (RWD) are collected in settings other than clinical trials and can be obtained through healthcare organizations’ electronic medical records (EMRs), insurance claims data, patient-reported outcomes (PROs), and mobile health technology. Using RWD has excellent potential for assessing treatment effectiveness in oncology, as such usage can improve patient quality of life and help develop novel treatments. The most important advantage of using RWD in oncology is that it can provide “real patient” data that clinical trials cannot capture. The Japanese healthcare system is characterized by the widespread use of EMRs. For example, the overall penetration of EMRs in Japan in 2020 was 57.2%, with an exceptionally high penetration rate of 91.2% among medical institutions with more than 400 beds [1]. Consequently, several clinical studies have also been conducted using medical records [2,3]. Japan has a universal health insurance system, ensuring that standard treatments are widely accessible to the population. However, only a few international observational studies have utilized Japanese electronic medical records, and there has been no integration of data with overseas facilities. Japan’s active involvement in observational oncology research will enable more comprehensive studies.

Recently, the Observational Medical Outcomes Partnership Common Data Model (OMOP CDM) [4], published by Observational Health Data Sciences and Informatics (OHDSI) [5], has become widely used in conducting observational studies. The OMOP CDM is a standard data model specifically designed for observational research, allowing EMR data into a format that can be exploited using the analytic tools and methods provided by the OHDSI.

The OMOP CDM is a powerful platform for integrating medical data from around the world, facilitating collaboration in observational research. Overseas, there have been initiatives to convert electronic medical record data to the OMOP CDM. In addition, in Japan, the project “Initiatives for the Creation of Real-World Evidence” (commonly known as Rinchu Net) conducted a survey on the OMOP CDM to facilitate participation in international research [6].

To date, there have been no studies on converting EMR data to the OMOP CDM in Japan, and its feasibility has not yet been fully verified. Therefore, it is necessary to construct an OMOP-based environment for technical verification to evaluate if EMR data can be converted to an OMOP CDM and used with OHDSI’s analysis tool (ATLAS) [7]. The aim of this study was to evaluate the feasibility of converting a representative Japanese oncology EMR data to the OMOP CDM standard. Specifically, we (1) assessed the fraction of source terms mapped to the OMOP CDM terms, (2) compared local raw datasets with the OMOP CDM standards, and (3) conducted comprehensive quality assessment using the Data Quality Dashboard (DQD) [8].

## Methods

### Dataset

This study was approved by the National Cancer Center Institutional Review Board of the National Cancer Center, Tokyo, Japan (Research Project number 2020-418). The source data was first accessed on 17th November 2023 and were extracted from the EMR system of the National Cancer Center Hospital East, [Chiba], Japan and covered 8,447 patients with breast cancer from January 2015 to November 2023. After excluding those whom the researchers deemed inappropriate such as the patients who participated in the clinical trial, a total of 8,387 patients were included in the study. The source data were in the TXT file format containing clinical information, including dates and timestamps, extracted from all inpatient, outpatient, and emergency visits. Source data included patient profile information, death records, visit details (inpatient and outpatient), disease names, drug therapies employed, specimen testing, vital signs, physical findings, medical and surgical histories, surgeries, radiation treatment, and physiological tests. Patient IDs and names, which could potentially identify patients, were replaced by research IDs specific to this study, and birthdates were adjusted to display years and months, with all data modified to reflect the first of the month. Patient location information was restricted to prefectures. Although the above steps were taken and the research was conducted in such a way that individuals could not be identified, access to Patient ID was still available from the Research ID in case a patient complaint or source data needed to be verified.

Table 1 presents the dataset characteristics.

**Table 1.**
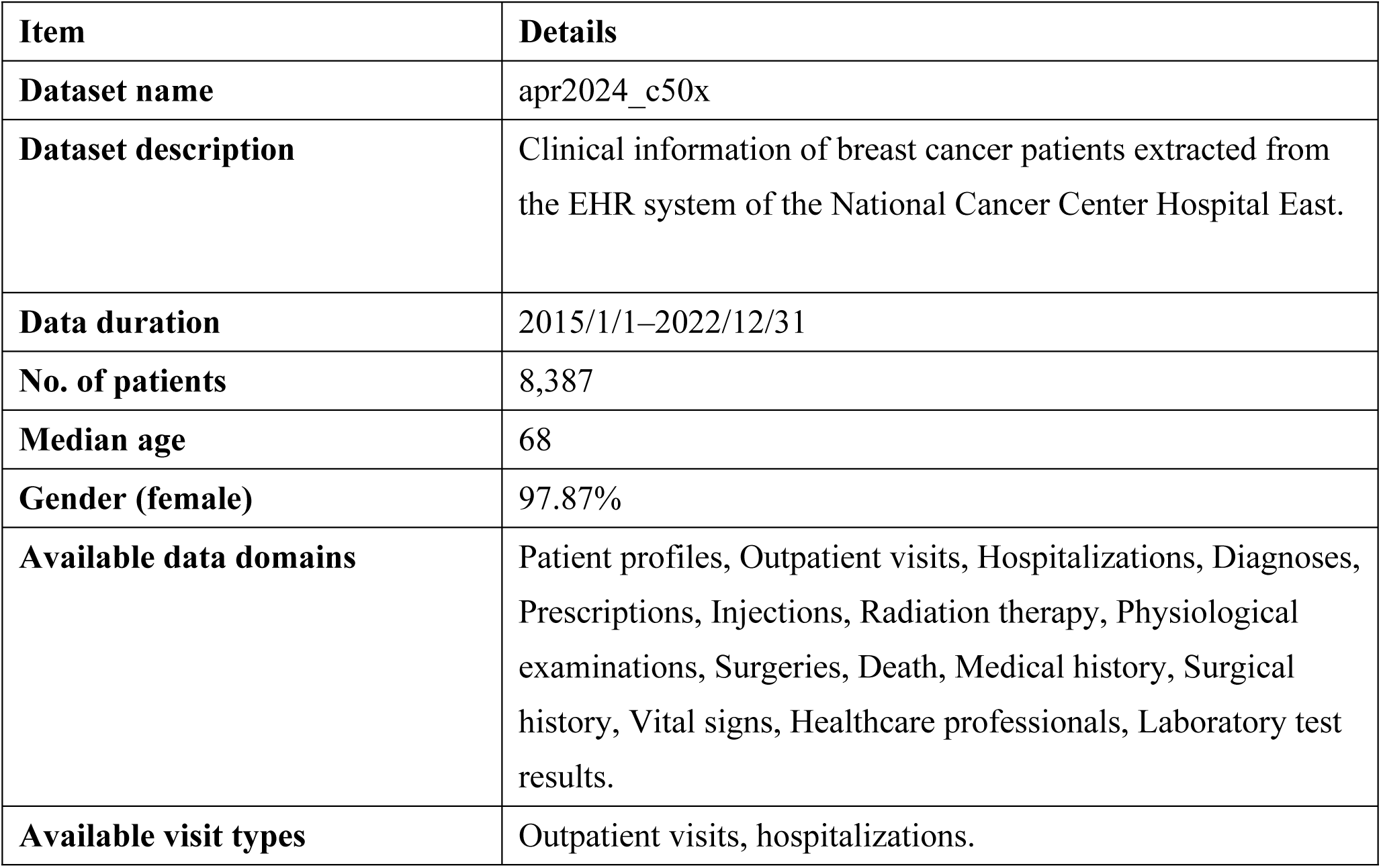
Dataset overview.

### OMOP CDM

The OMOP CDM is a patient-centric model maintained by the OHDSI community that allows patient data to be stored across different domains. Thirty-nine tables (version 5.4) were structured into various domains including clinical, health systems, health economics, metadata, vocabulary, and derived elements. Tables for each domain and details of the CDM rules for entering data under those tables can be obtained by referring to the online OMOP CDM documentation. In this study, we implemented OMOP CDM version 5.3.1. Specifically, we entered the data into 16 clinical tables, including the care site, condition occurrence, death, drug exposure, drug strength, location, measurement, observation, observation period, person, procedure occurrence, provider, and visit occurrence.

### Conversion of EMR data into an OMOP CDM

Two main tasks are required to convert EMR data into an OMOP CDM: 1) standardization of vocabulary and 2) standardization of the data structure. The details of this process are described below.

#### 1) Vocabulary standardization

When converting source data into an OMOP CDM, it is necessary to convert the source vocabulary used in the EMR into a standard vocabulary that the OMOP CDM can analyze. RxNorm and RxNorm-Extension are examples of drugs, and LOINC and SNOMED are examples of tests. Please refer to the Book of OHDSI [9] and ATHENA [10] for the standard vocabulary published by the OHDSI.

To convert the Japanese EMR vocabulary to a standard vocabulary, we first obtained the disease name master, specimen test result master, drug master, radiology master, physiological test master, and surgery master from the EMRs. Next, we checked the frequency of each vocabulary item by referring to the EMR data and prioritized mapping the most frequent items first. This exercise led to mapping of up to 90% of the source data. We focused on structured data and did not perform tasks, such as breaking down sentences into vocabulary. Next, mapping was performed using a standard vocabulary following the OHDSI procedure, with two main mapping scenarios. In Scenario 1, the source vocabulary had standard concept mapping in ATHENA. For example, in Japanese EMRs, illness names are often expressed using ICD-10, which is already registered as a source term in ATHENA, and can be quickly processed into the standard vocabulary. In Scenario 2, the source vocabulary lacked standard concept mappings in ATHENA. Although the Ministry of Health, Labour, and Welfare in Japan has published standard vocabulary [1], these do not have standard concept mappings in ATHENA, requiring the creation of unique mappings. In our hospital, we used Scenario 1 for disease name registration and Scenario 2 for other domains.

#### 2) Standardization of data structure

Converting source data to the OMOP CDM requires mapping the source data attributes to the correct columns in the appropriate OMOP CDM tables. The OMOP CDM schema, consisting of 39 tables, is available on the OHDSI GitHub repository [4]. When constructing the schema, we referred to the Japanese environmental documentation at OHDSI Japan [11]. We did not convert the cause of death in this study because it could potentially lead to patient identification. The following tasks were performed when mapping the source data to the OMOP CDM tables:

- Changed patient chart IDs to research IDs, converted birthdates to year and month format, and standardized all dates to the first of the month.
- Identified and addressed data required in the OMOP CDM that were not present in Japanese charts. We verified this conversion by referring to a previous study [12].
- Evaluated the mapping coverage and calculated the percentage of source terms that could be expressed as concepts in the OMOP CDM format.
- Used the Data Quality Dashboard [8] to perform comprehensive validation checks on the suitability, completeness, and validity of data in the CDM dataset.

### Ethic Statement

This study (Project Number 2020-418) was conducted following the ethical principles outlined in the World Medical Association Declaration of Helsinki on Ethical Principles for Medical Research Involving Human Subjects. The need for informed consent was waived due to the retrospective nature of the study. This study was reviewed and approved by the Ethics Review Board of the National Cancer Center of Japan.

## Results

### EMR to OMOP CDM Conversion

We used data from the National Cancer Center Hospital East. The raw data were representative of 8,447 patients, but the number of patients converted to the OMOP CDM standard was 8,387. The reduction in the number of patients was due to the exclusion of patients who had participated in clinical trials and fictitious patients. The patients included the study provided clinical information on diagnosis, laboratory tests, visits, medications, observations, surgery, and death.

#### 1) Vocabulary standardization

In the vocabulary standardization phase, 3,697 unique disease names, 987 unique specimen test result terms, and 1,144 unique drug terms for prescription and injection were mapped to the OMOP CDM standard. The other terms and mapping ratios are shown in Fig 1. Our hospital uses ICD-10 for disease name data, and mapping to standard terms had already been registered in ATHENA; consequently, all terms could be mapped to standard terms. Only 32 surgical terms were identified.

**Fig 1.**
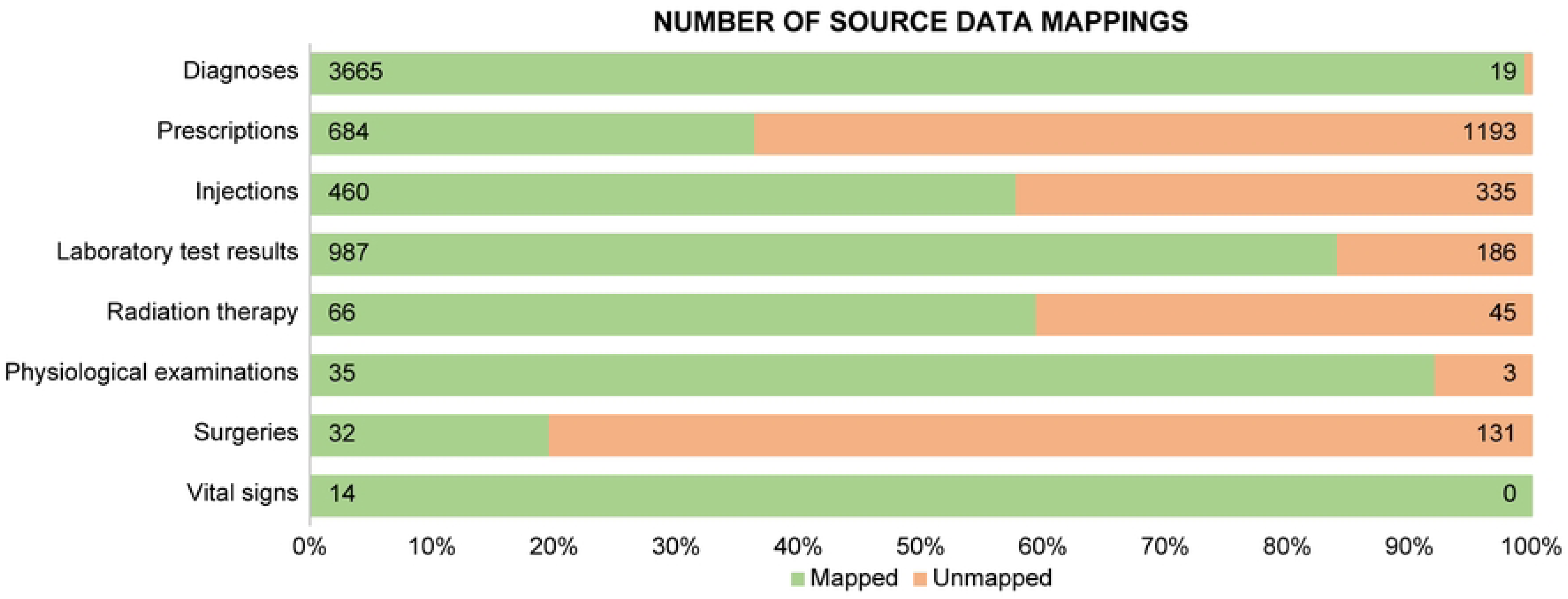
Conversion rate of vocabulary used in the hospital to standard vocabulary.

#### 2) Standardization of data structure

In line with previous studies evaluating the feasibility of the OMOP CDM, information on key demographic and clinical factors was extracted and compared between the EMR and OMOP CDM data. The source data from the EMR were converted into tables in the OMOP CDM, as shown in Fig 2. The success rate of the conversion was confirmed; the results are displayed in Fig 3.

**Fig 2.**
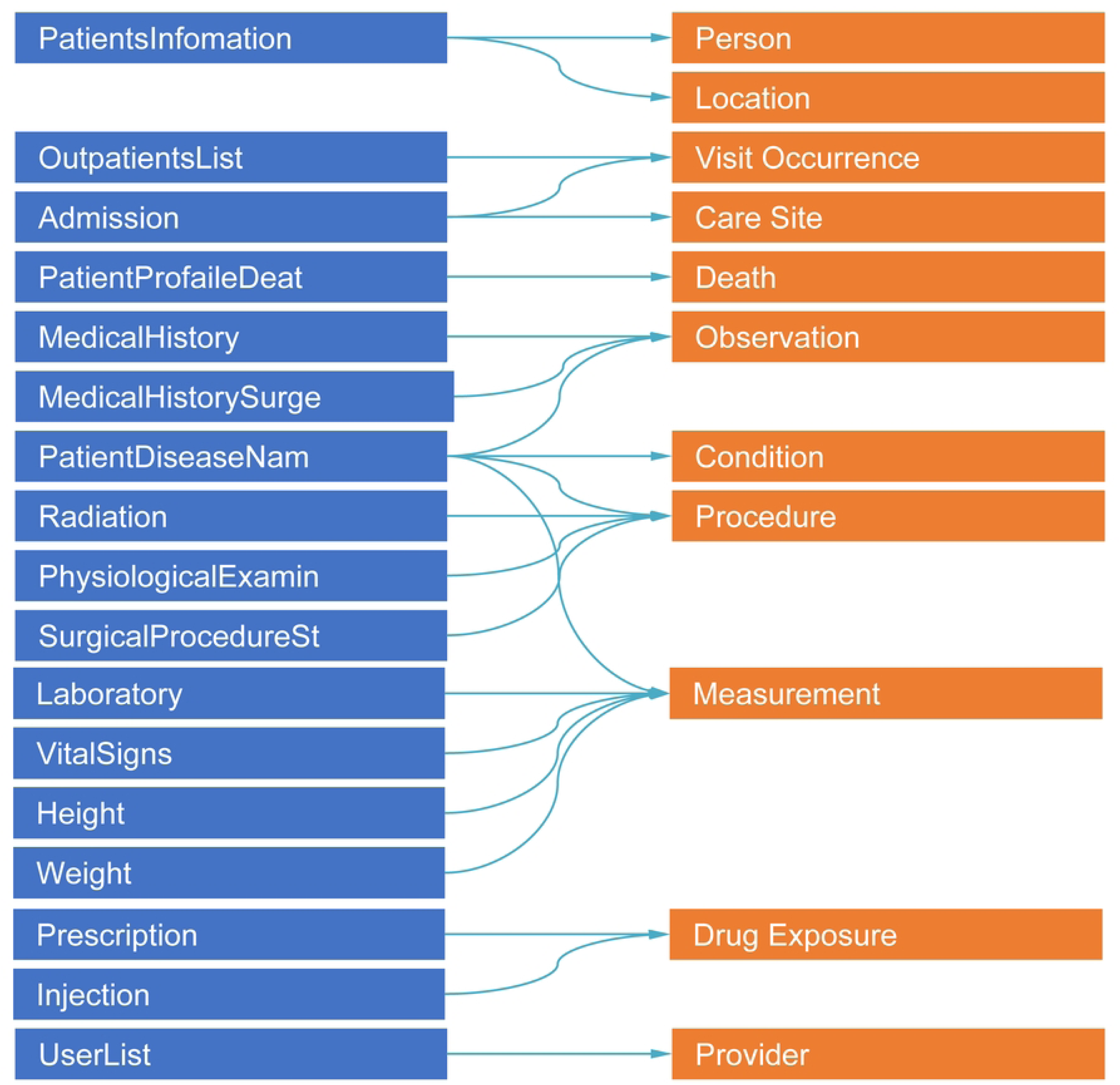
Analysis of the conversion of the database of the hospital information system into an OMOP structure.

**Fig 3.**
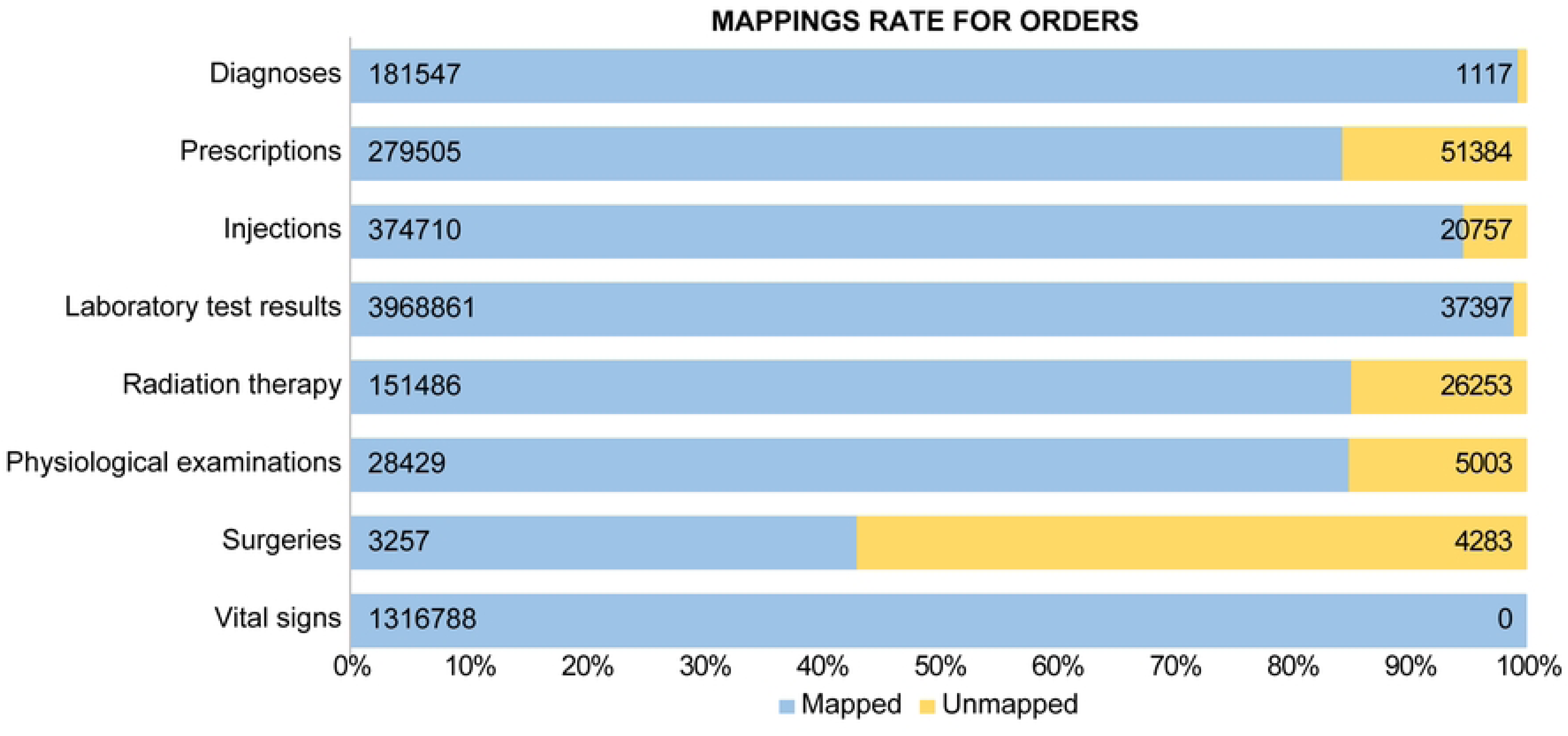
Comparison of electronic medical record structures with OMOP structures.

We assessed the quality of the converted dataset using the DQD, an open-source R package developed by the OHDSI community. A total of 1,991 validation checks were performed based on the data validity, suitability, and completeness of OMOP CDM data. The results of the DQD are presented in Fig 4. The list of checks used by the DQD can be found in the GitHub repository [8]. Consequently, 24 of the 1,991 validation checks performed in this study were either FAIL or ERROR. Upon examining the details of these 24 checks, we found that 11 affected plausibility and 13 affected completeness. When analyzed by table, the errors were distributed as follows: 2 in CONDITION_OCCURRENCE, 2 in DEATH, 1 in DEVICE_EXPOSURE, 4 in DRUG_EXPOSURE, 10 in MEASUREMENT, 1 in OBSERVATION, 1 in PAYER_PLAN_PERIOD, and 3 in PROCEDURE_OCCURRENCE. We were able to confirm the reasons for these findings (data not shown).

**Fig 4.**
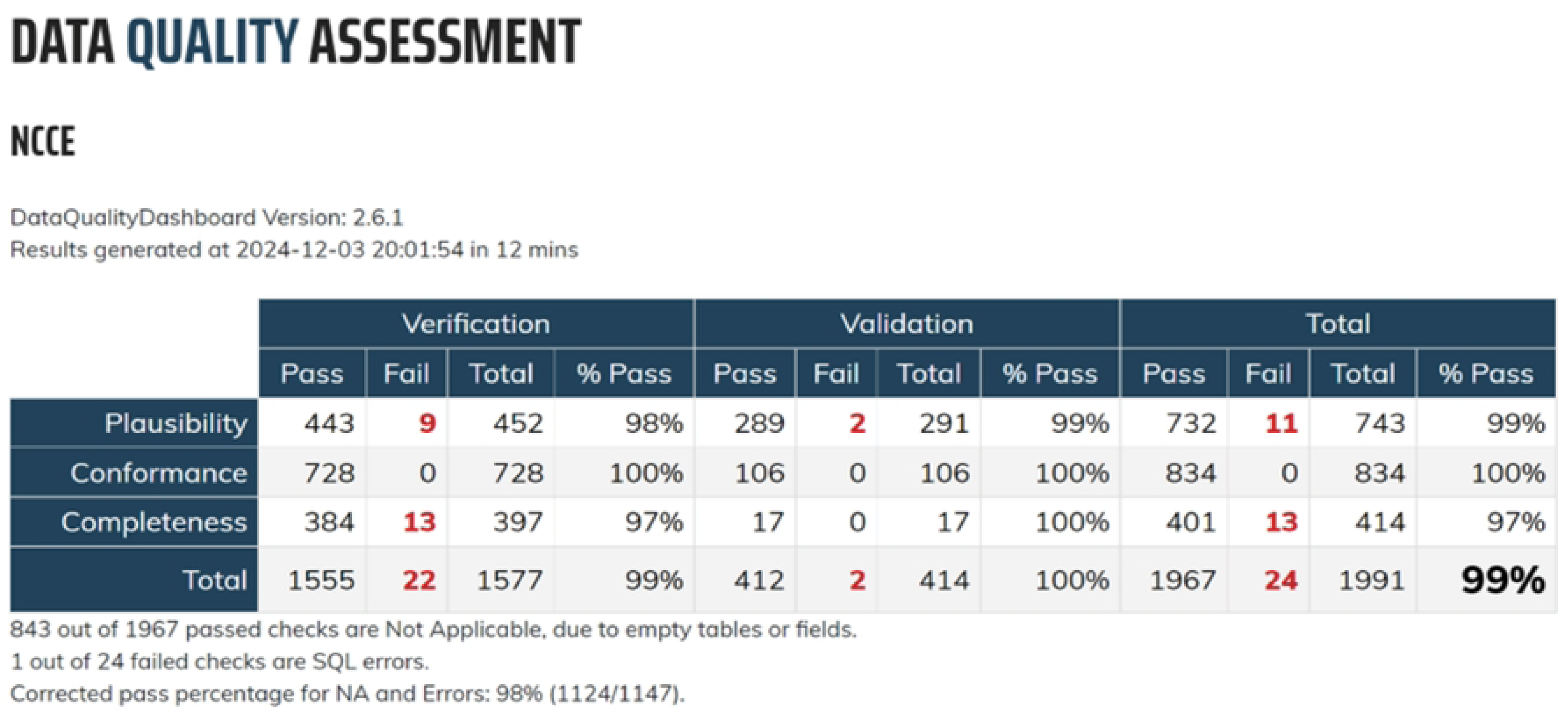
Quality Assessment using Data Quality Dashboards.

## Discussion

### Conversion of EMR to the OMOP CDM

In this study, we converted EMR data from Japanese medical institutions into the OMOP CDM, a process that has been rarely reported in Japan and is considered highly valuable. Here, we discuss the advantages and challenges associated with OMOP use in Japan.

#### Vocabulary standardization

The source terms used in the EMR were successfully mapped to standard terms in the OMOP CDM. However, complex concepts, such as surgery, could not be mapped to any distinct term. Clinical test terms were easier to map to SNOMED than to LOINC, with most results mapped to SNOMED. While LOINC helps in understanding specific clinical tests performed, it was not easy to determine which LOINC terms corresponded to the Japanese test items. In contrast, SNOMED provided a more general understanding of test procedures, making mapping easier. ICD-10 codes, the commonly used disease coding system in Japan, were already mapped to standard terms in ATHENA, making it easy to convert to the standard terms. However, for other vocabularies, mapping from source terms to standard codes was required, and processing the 11,869 terms required considerable effort. In Japan, standard codes are defined by the government, and although these codes are easy to use, they are not mapped to the standard terms defined by the OHDSI. We matched standard terms directly to the source terms used in the hospital, although this imposed considerable workload. If Japanese standard codes become widely used and mapping information to standard terms of the OHDSI becomes available in ATHENA, converting EMR data to the OMOP CDM could become easier. However, converting regimen information, essential for managing cancer treatments, to OMOP was impossible. We suggest that future studies should enter the regimen name into the OMOP CDM, which will provide an overview of changes in a patient’s treatment. To store regimen information in the OMOP CDM, the database required expansion, for example, by using the Oncology Extension [13]. This issue should be addressed in the near future.

### Standardization of data structure

Next, using the standardized vocabulary, we created an SQL program to convert the EMR source data into an OMOP CDM. Before creating the program, we reviewed the source data and resolved certain issues (for example, we clarified the EMR data items required for the OMOP CDM). Overall, we successfully standardized more than 80% of the domains, excluding surgery. In particular, the domains such as disease name, Laboratory test results, physiological testing, and vital signs also showed successful vocabulary standardization and good clinical data standardization. Interestingly, data structures were standardized even in domains where vocabulary standardization was not fully achieved (e.g., drugs and radiation), owing to the selection of the vocabulary-standardization target. In the OHDSI methodology, conversion is typically performed on frequently used vocabulary covering up to 80% of the data. In our study, up to 90% conversion was achieved on the vocabulary for drugs and radiation. As a result, the clinical data were successfully standardized. This approach allows for sufficient standardization while minimizing the burden (workload) on the operator. However, oncology-specific items, such as drugs and tests, must be selected and converted, even if their occurrence is low. Terminology mapping for the surgical data was unsuccessful; therefore, clinical data standardization was not achieved for this domain. These results revealed the variations in how surgeries are recorded in the system. In addition, we encountered challenges with missing or unavailable data in certain OMOP CDM columns. Most of the data were available from medical records, but data pertaining to race and ethnicity were not generally available in the Japanese EMRs. Although this information is often incorporated into clinical trials or research, it was not available for all patients who visited the hospital. Moreover, vital signs such as body temperature, heart rate, respiratory rate, and blood pressure were not recorded with specific units. This information could be gathered only visually from the EMR screen; therefore, we had to supplement the units with data. Similarly, data on height, weight, and some clinical testing locations were old, and the measurements were unclear. Other issues included unclear dates for certain medical tests and missing dates. Results without a measurement date were excluded from the conversion process.

The results of the DQD showed 99% conformity for the test items, indicating overall good quality. A detailed review of the 24 FAIL or ERROR cases revealed errors related to sex, missing source data in the DEATH table, errors in the unit conversion of clinical test results, incorrect drug dosage conversions, and unregistered source data. These issues can be addressed by modifying the conversion program or reviewing the source-to-concept map. In some cases, further review of source data may be needed, such as addressing inconsistencies between disease names and sex. One possible measure to improve these results is to prepare a correspondence table between the Japanese terms and OHDSI standard terms. Currently, ATHENA does not link Japanese terms to the OHDSI standard terms; therefore, each institute must create their own mapping table. However, Japan has launched a national project to standardize the secondary use of EMR data, and medical institutions are beginning to adopt government-defined standard terminology. [14] [15]. In the future, as Japanese standard terms become linked to OHDSI standard terminology, the use of OMOP in Japan will likely improve, enabling OHDSI research.

Vocabulary mapping is a resource- and knowledge-intensive task. To reduce these burdens, technologies such as AI are currently being explored. For example, the Kimura group (Ehime University, Ehime, Japan) has developed a semi-automated mapping process using a large-scale language model (LLM) to incorporate Japanese drug codes into the OHDSI project [16]. These methods are expected to make Japanese standard terminology more accessible for OHDSI research. It is also expected that an operational system will be established to manage these terms and register them properly in repositories such as ATHENA. The most significant advantage of using the OMOP CDM is the ability to use standardized tools and network research methods. Network research, actively conducted at the OHDSI, allows medical institutions worldwide to integrate and analyze data converted into the OMOP CDM, generating valuable evidence. A major benefit of network studies is the clustering of cases. Recent initiatives, such as efforts to improve interoperability through Fast Healthcare Interoperability Resources (FHIR) attempts to link OMOP data with FHIR, have allowed the collection and conversion of data from medical institutions into the OMOP for further analysis. This collaboration between the FHIR and OMOP can potentially create greater consistency between source data and evidence generation. In Japan, the development of an electronic medical record sharing service is underway, and the FHIR will be used for this purpose. If this service can enable analysis using the OMOP CDM within this service, it could create a large medical network for international research. By introducing this knowledge and infrastructure in Japan, large-scale research utilizing the existing medical data may become possible. Currently, efforts to convert data to the OMOP model are being focused on research use. However, initiatives to use the OMOP CDM for hospital management are also underway. For example, Park et al. demonstrated the usefulness of CDM for healthcare process mining and proved its usefulness.[17] Healthcare process mining involves the analysis of events that occur in multiple processes, such as inpatient, outpatient, and emergency room visits, as well as patient transfers, to derive process-related insights. The use of OMOP CDM for healthcare process mining has proven effective as a data source for analyzing healthcare processes. It also enables the application of the same analysis method across different institutions. Efforts to improve the performance are crucial when handling a large volume of data. Kang et al. have been working on converting a relational database into a graph database schema and have reported that these efforts have considerably improved data creation and querying capabilities.[18] Many resources are required to convert the raw data into the OMOP CDM. However, only a few such cases have been reported in Japan. As the number of cases increases, challenges related to the data stored in the EMR and knowledge required for the conversion will become more apparent, leading to further advances in this area.

## Conclusions

This feasibility study revealed that Japanese EMRs can be appropriately converted into the OMOP CDM. It also demonstrates the urgent need to link Japanese vocabulary with the OHDSI standard vocabulary for more efficient conversion. These tasks still require significant resources but must be carried out in cooperation with the government and other organizations. Furthermore, as more cases are reported, the potential for international research and healthcare process analysis using OMOP is bound to grow.

### Clinical Relevance Statement

Using the OMOP CDM, it is possible to standardize data from various sources into an international format. This study enables a global comparison of real-world treatment conditions in the field of oncology. This approach will provide essential insights into the field of oncology and have a positive effect on future treatment plans. Japan has a universal health insurance system, and standard treatment strategies are often adopted. By comparing these data with the expected results of clinical trials, real-world situations can be better understood.

## Data Availability

The datasets generated and analyzed in this study are not publicly available as consent to provide data to non-researchers has not been obtained but are available from the corresponding author upon reasonable request.

## Acknowledgments

The authors thank IQVIA Solutions and Fujitsu for providing valuable technical assistance throughout EMR data transformation. The authors would also like to thank the Japan Agency for Medical Research and Development (AMED) and Rinchu-net for government projects.

## Notes

### Competing Interest Statement

The authors have declared no competing interest.

### Funding Statement

Yes

### Author Declarations

National Cancer Center Institutional Review Board 2020-418

